# Associations of weather and air pollution with objective physical activity and sedentary time before and after bariatric surgery: a secondary analysis of a prospective cohort study

**DOI:** 10.1101/2023.03.22.23287589

**Authors:** Aurélie Baillot, Paquito Bernard, Nejm Eddine Jmii, J. Graham Thomas, Leah M. Schumacher, Pavlos K. Papasavas, Sivamainthan Vithiananthan, Daniel Jones, Dale S. Bond

**Author notes:** CORRESPONDING AUTHOR: Aurélie Baillot, Université du Québec en Outaouais, 283 Boul. Alexandre-Taché, Gatineau, Québec, Canada, J8X 3X7.

## Abstract

**Background:** Most metabolic and bariatric surgery (MBS) patients perform too little moderate-to-vigorous intensity physical activity (MVPA) and too much sedentary time (ST). Identifying factors that influence MVPA and ST in MBS patients is necessary to inform the development of interventions to target these behaviors. Research has focused on individual-level factors and neglected those related to the physical environment (e.g., weather and pollution). These factors may be especially important considering rapid climate change and emerging data that suggest adverse effects of weather and pollution on physical activity are more severe in people with obesity.

**Objectives:** To examine the associations of weather (maximal, average and Wet Bulb Globe Temperatures), and air pollution indices (air quality index [AQI]) with daily physical activity (PA) of both light (LPA) and MVPA and ST before and after MBS.

**Methods:** Participants (n=77) wore an accelerometer at pre- and 3, 6, and 12-months post-MBS to assess LPA/MVPA/ST (min/d). These data were combined with participants’ local (Boston, MA or Providence, RI, USA) daily weather and AQI data (extracted from federal weather and environmental websites).

**Results:** Multilevel generalized additive models showed inverted U-shaped associations between weather indices and MVPA (R^2^≥ .63, p<.001), with a marked reduction in MVPA for daily maximal temperatures ≥20°C. Sensitivity analysis showed a less marked decrease of MVPA (min/d) during higher temperatures after versus before MBS. Both MVPA before and after MBS (R^2^=0.64, p<.001) and ST before MBS (R^2^=0.395; p≤.05) were negatively impacted by higher AQI levels.

**Discussion:** This study is the first to show that weather and air pollution indices are related to variability in activity behaviors, particularly MVPA, during pre- and post-MBS. Weather/environmental conditions should be considered in MVPA prescription/strategies for MBS patients, especially in the context of climate change.

## Introduction

Physical inactivity and excessive sedentary time are major concerns in the context of metabolic and bariatric surgery (MBS). While regular physical activity can optimize the short- and long-term benefits of MBS^1^, many MBS patients are insufficiently active and also spend a large part of their waking time sedentary before and after MBS^2,3^. Thus, promoting adoption and maintenance of higher physical activity levels is an important facet of multidisciplinary care of the MBS patient^4^.

Identifying factors that influence physical activity and sedentary behaviors in MBS patients is essential to inform the development of effective behavioral interventions to promote a more active lifestyle. However, most research has focused on individual-level factors and largely neglected environmental factors, especially those related to the physical environment. According to the socio-ecological model^5^, factors that influence behavior should be considered from multiple levels of influence including environmental. Moreover, the environmental level of influence on behavior is increasingly important in the context of global climate change and global warming.

Two environmental factors that may influence physical activity and sedentary patterns of adults generally and MBS patients specifically are shifts in weather patterns and amount of air pollution^6^. Recent reviews of studies involving participants across multiple countries and continents show that total physical activity volume is consistently greater during higher outdoor temperatures whereas sedentary behavior is greater during lower outdoor temperatures^7-9^. Studies also show that Wet Bulb Globe Temperature (WBGT), a more sophisticated indicator of outdoor temperature that combines the effects of radiation, humidity, temperature and wind speed on the perception of temperature, is negatively associated with daily steps^10^, active transport^11^, and outdoor occupational related physical activity^12^.

Similarly, cross-sectional and longitudinal studies indicate that increased air pollution is associated with lower physical activity volume^13^. One US study in particular showed that an increase of fine particle matter (i.e., respirable particle with 2.5 microns in diameter or less) was related to a 16-35% increase in the odds of leisure physical inactivity among adults^14^. Conversely, a study conducted in China found that an increase of one standard deviation in air pollution, measured via the Air Quality Index (AQI, i.e., index combining carbon monoxide, nitrogen dioxide, ozone, sulphur dioxide, particulate matter measures), was related to a 7.3 hour increase in weekly self-reported sedentary time^15^.

Among adults with obesity, studies report that weather conditions can be a barrier to physical activity^16,17^. Both higher (snow) precipitation and temperature are associated with lower physical activity levels among adults with severe obesity^18,19^. Additionally, growing research suggests that the adverse effects of weather pattern shifts and air pollution on physical activity may be more severe in adults with obesity. Findings of a US study suggest that adults with obesity are more vulnerable to heat stress, and that this vulnerability contributes to larger reductions in physical activity. Specifically, results showed a non-linear inverse U shape association between daily maximal temperature and leisure-time physical activity volume, where physical activity participation decreased in temperatures that exceeded 35°C. This decrease was seven times higher in adults with overweight or obesity in comparison to adults with Body Mass Index (BMI) < 25 kg.m^2 19^.

However, to our knowledge no study has investigated potential associations of weather and air pollution with physical activity and sedentary time in adults with obesity before and after MBS. It is possible that physiological changes after MBS could help reduce the degree to which outdoor temperatures and air pollution are a barrier to physical activity via increased tolerance to higher outdoor temperatures (via reduced adipose tissue and improved thermoregulation) and air pollution (via increased respiratory function). This is a particularly important knowledge gap in light of current and future climate changes. Heat waves are occurring more often during the last decade (from 2 per year in 1960 to 6 per year in 2020)^20^. The sixth Intergovernmental Panel on Climate Change report concluded that (extreme) temperatures are expected to increase in North and Central America, as well as the number, the frequency and the severity of natural disasters (e.g. heat waves, intense precipitation)^21^ causing health and behavioral impacts^22^.

In light of the limited evidence, coupling with the climate change context, our study aimed to examine the associations of objective daily weather (maximal and averaged temperature, WBGT) and air pollution indices (AQI) with device-measured daily time spent in physical activity (light, and moderate-to-vigorous intensities) and sedentary behaviors before and after MBS. We hypothesized that higher daily weather temperatures, WBGT and AQI levels would be associated with less daily time spent in physical activity, especially MVPA, and more daily time spent in sedentary behaviors.

## Methods

### Participants

This study involves analysis of data from a prospective cohort study that was designed to evaluate multiple behavioral and psychosocial predictors of outcomes before and during the first year after bariatric surgery. To be initially eligible, participants had to have a body mass index (BMI) ≥ 35 kg/m^2^, be ≥ 21 years old, and be scheduled to undergo Roux-en-Y gastric bypass (RYGB) or sleeve gastrectomy (SG) at one of two university-based hospitals in the Northeastern United States. Participants were excluded if they were receiving weight management treatment outside the context of standard surgical care, or reported presence of a condition (e.g., uncontrolled severe mental illness) or factors (e.g., plans to geographically relocate) that could preclude adherence to the study protocol. A total of 92 participants consented to participate at baseline, 77 of whom provided accelerometry data with ≥7 days at baseline. Then, 60 (78%), 54 (70%), and 44 (57%) participants, providing accelerometry data with ≥7 days were included in analysis at follow-up, at 3-, 6-, and 12-month post-surgical visits, respectively (Supplemental Figure 1 for flow chart).

### Procedure

All aspects of the parent study protocol that are relevant to the present analyses are described below; the full study protocol is published elsewhere^23^. This study is reported in accordance with the guidelines for reporting observational studies (Strengthening the Reporting of Observational Studies in Epidemiology (STROBE)) (Supplemental Table 1).

Participants were recruited individually on a rolling basis between May 2016 and April 2018 at a regularly scheduled clinic visit occurring 3-8 weeks before their scheduled surgery date. Participants deemed initially eligible after telephone screening complete an in-person screen/baseline assessment at the bariatric clinic or affiliated research center. During this initial visit, participants provided informed consent, had their height and weight measured, completed a demographic questionnaire, and were provided with an accelerometer to complete 10 days of activity monitoring prior to surgery. Participants wore the accelerometer again at 3, 6, and 12 months after surgery. For the current study, daily accelerometer data from participants were combined with daily weather data using each participant’s city address/location and accelerometer wear time dates. Measures and data sources are detailed below. The parent study was approved by the institutional review boards of The Miriam Hospital (TMH) in Providence, RI, USA and Beth Israel Deaconess Medical center (BIDMC) in Boston, MA. The study was registered at www.clinicaltrials.gov (NCT02777177).

### Measures

#### Objectively-measured physical activity and sedentary behaviors

Participants’ daily minutes spent in physical activity and sedentary behaviors were recorded using an ActiGraph GT9X Link wrist-worn triaxial accelerometer. Valid wear time was defined as ≥ 7 days with ≥ 10 hours of wear per day. Sleep periods were identified and removed from analysis using the Cole-Kripke and Tudor-Locke algorithms within the ActiLife Version 6.13.3 software (ActiGraph, LLC, Pensacola, FL, USA). Nonwear periods, defined as ≥90 minutes without movement using vector magnitude counts and with allowance of interruptions of ≤2 minutes of nonzero counts, were also identified and removed ^24,25^. Vector magnitude counts per minute thresholds that have been shown to minimize the mean difference between estimates of sedentary time (ST) and MVPA when using wrist-versus hip-based ActiGraph accelerometers were used to estimate minutes per day spent sedentary (< 2000 counts per minute) and in light-intensity physical activity (≥2000 and <7500 counts per minute) and MVPA (≥ 7500 counts per minute)^26^.

### Independent variables: weather indices and air pollution data

Physical activity and sedentary daily data across assessments were combined with local daily weather data according to participants city localization (Boston or Providence) and accelerometer wear time dates. Averaged and maximal temperatures (C°), precipitation and snow fall (cm) were obtained from the USW00014739 and USW00014765 weather stations, which were the closest to Boston and Providence. AQI data were acquired from monitoring stations which are present in core-based statistical areas of both Boston-Cambridge-Newton and Providence-Warwick. AQI is an index combining carbon monoxide, nitrogen dioxide, ozone, sulphur dioxide, and particulate matter measures ^27^. Weather data and AQI were extracted from the websites of the National Center of Environment Information (NCEI) of the National Oceanic and Atmospheric Administration (NOAA), and the Environmental Protection Agency, respectively. Higher AQI value are associated with a greater level of air pollution. An AQI >50, >100, > 200, and > 300 were considered as moderate, unhealthy for sensitive groups, unhealthy, very unhealthy, and hazardous, respectively^28^. There is a high health risk to perform MVPA for unfit or non-acclimatized adults with WBGT > 25.7°C (78.1°F)^29^.

### Covariates

Sociodemographic covariates included gender, age, ethnicity, level of education, and body mass index (calculated with weight and height measured).

### Statistical analysis

A set of t-tests was carried out to compare study participants recruited in Boston or Providence. To investigate the association between environmental and movement related data, the shape of relationships (linear versus non-linear) was first investigated^7,30,31^. Consequently, multilevel linear and generalized additive models (GAM)^32^ were carried out for each tested association. Participants were nested within their respective recruitment center. The GAM is an extension of the generalized linear model in that one or more predictors may be specified using a smooth function. GAM is a nonparametric model that allows nonlinear relationships to be modeled with flexibility without specifying the nonlinear functional form ^31^. Non-linearity in GAM was based on the effective degrees of freedom (edf) of the smothing terms, with edf >3 indicating a non-linearity shape. These models included movement related variables (MVPA, LPA, sedentary time) as dependent variables, and time varying covariates (days number, season, weekend day (Yes/No), precipitation, snowfall). To take time autocorrelation into account, lag-1 of dependent and weather variables were also included in models. A cubic spline smoother was used to fit GAMs. Both models were compared using Akaike’s Information Criterion (AIC), with smaller values for the information criterion indexes indicating a better model^33^. Then, the selected model (linear or non-linear) was executed again by including socio-demographic covariables (i.e., age, gender, BMI). If a statistically significant association was found in GAMs, a 2-dimensional plot was produced. The packages mgcv, tidyverse, lmer, gtsummary, hydroTSM and weathermetrics form the statistical software R (v4.2) were used to prepare, conduct and visualise the models. The R syntax for main analyses are provided on the Open Sciences Framework (https://osf.io/34usq/). To examine the potential effect of MBS, a set of sensitivity analyses and visual examination of plots was performed^34^. All selected full models were separately carried out for pre- and post-surgery data.

## Results

### Population characteristics

Of the 77 participants included before the surgery, 86% were women, 48% identified as other than White ethnicity, 37% had earned a college or graduate degree and the mean age was 44.5±11.3 years. The average baseline BMI was 45.9±7.6 kg/m^2^. Detailed information on accelerometer data (LPA, MVPA, sedentary time), BMI and data collection season according to the 4 times assessments and seasons are presented in Supplemental Table 2.

### Weather indices and air pollution description

The physical activity and sedentary time assessments were performed in Providence and Boston, having had a mean temperature of 12.1±9.1C° (range: -15.0 to 31.7C°), a WBGT score of 48.5±15.4 (range: 5.0 to 77.0), and an AQI score of 49.6±17.5 (range: 21.0 to 151.0) between June 2016 and April 2019. The air quality during the accelerometer data collection was good (<50), moderate (50-100), and unhealthy (>100) for 62%, 35% and 3% of days, respectively. Also, all accelerometer data were collected during days with a WBGT <25.6C° (78.1°F), which is below the high-risk threshold to perform MVPA for unfit or non-acclimatized adults.

### Associations of weather indices and air pollution with MVPA

Non-linear models had a better fit index and showed a statistically significant association between tested weather indices, AQI and MVPA (Figure 1, and supplemental Table 3). Each model is plotted in Figure 1, the lines show the smoothed function and dash lines indicated the 95% confidence area. Average and maximal temperature were significantly associated with MVPA, p < 0.001, R^2^ = 0.64, and p < 0.001, R^2^ = 0.64, respectively. A visual examination of plots indicated an important reduction of daily minutes of MVPA after ∼13°C (∼55°F) and ∼ 20°C (∼68°F) for average and maximal daily temperatures, respectively. An inverted U-shaped was found for association between daily MVPA with WBGT (p < 0.001, R^2^ = 0.63). Highest MVPA level was observed between ∼ -7°C (∼20°F) and ∼14°C (∼57°F) WBGT, with a drastic reduction of MVPA below and above these values.

**Figure 1:**
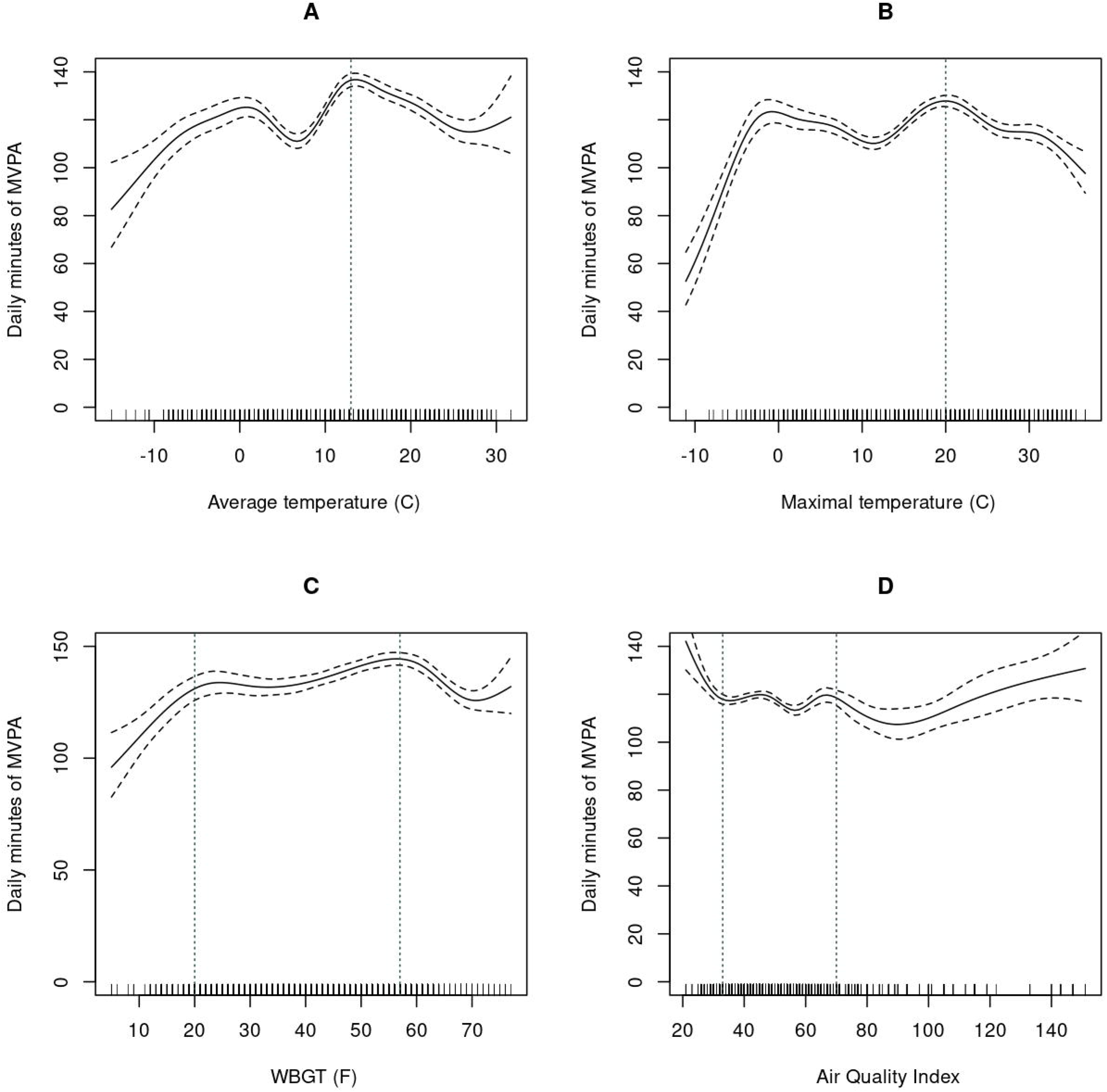
Associations of weather indices (average temperature, maximal temperature, WBGT) and air pollution (Air quality index) with moderate-vigorous intensity physical activity. Note: The lines show the smoothed function from GAM for daily minutes of MVPA, and the shaded area indicates the 95% confidence interval. Each model is adjusted for age, body mass index, gender, days number, season, weekend day (Yes/No), precipitation, snowfall, lag-1 of MVPA and weather variables. Vertical lines are placed based on visual observation.

Concerning AQI, a statistically significant association with MVPA (p < 0.001, R^2^ = 0.64) was observed with a non-linear complex shape. A decrease of daily time spent in MVPA was observed until ∼33 AQI score, followed by a monotonic curve, and an MVPA decrease after ∼70 AQI score.

### Associations of weather indices and air pollution with LPA

The shape of the associations of LPA with averaged temperature, maximal temperature, and WBGT were non-linear, but linear with AQI. No statistically significant associations were found between LPA and averaged temperature, maximal temperature, WBGT, AQI in multivariate models (Supplemental Table 4).

### Associations of weather indices and air pollution with sedentary time

Non-linear models were selected for tests involving sedentary behavior. No statistically significant associations of weather and air pollution outcomes with daily time spent in sedentary behaviors were found (Supplemental Table 5).

### Sensitivity analyses: before and after MBS

The association shapes for MVPA were globally similar between pre- and post-MBS (Figure 2, and supplemental Table 6). However, a visual examination of plots indicated that a more drastic reduction in daily minutes of MVPA appears before MBS for average temperature at ∼ 20°C and for WBGT at ∼8°C (∼ 47°F) compared to after MBS. No significant associations were observed between LPA and weather outcomes in pre and post-MBS (Supplemental Table 7). Additionally, a positive linear statistically significant association was found between daily time spent in sedentary behaviors and AQI before MBS (R^2^=0.395; p≤.05) (Figure 3, and supplemental Table 8).

**Figure 2:**
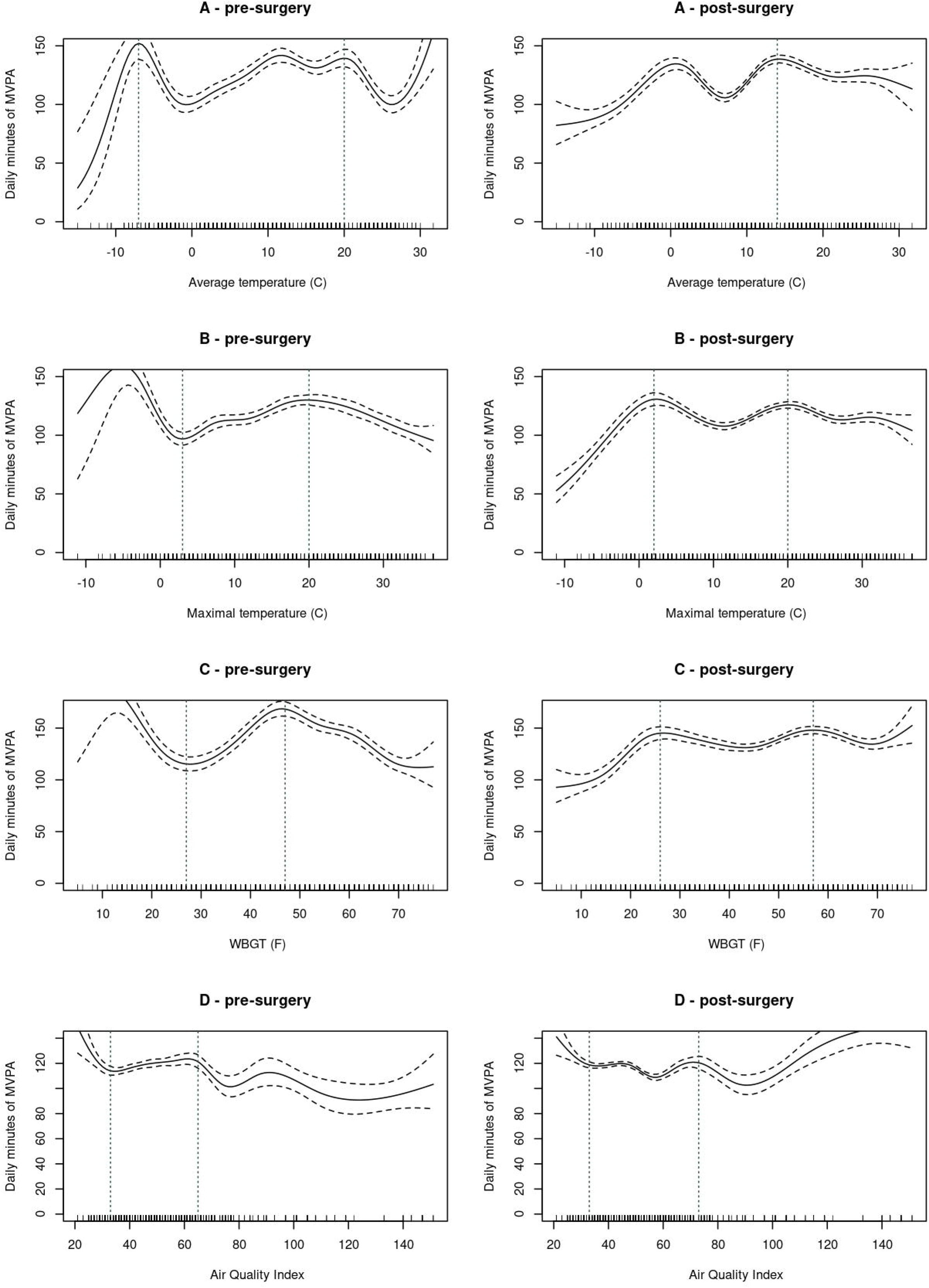
Associations of weather indices (average temperature, maximal temperature, WBGT) and air pollution (Air quality index) with moderate-vigorous intensity physical activity before and after MBS. Note: The lines show the smoothed function from GAM for daily minutes of MVPA, and the shaded area indicates the 95% confidence interval. Each model is adjusted for age, body mass index, gender, days number, season, weekend day (Yes/No), precipitation, snowfall, lag-1 of MVPA and weather variables. Vertical lines are placed based on visual observation.

**Figure 3:**
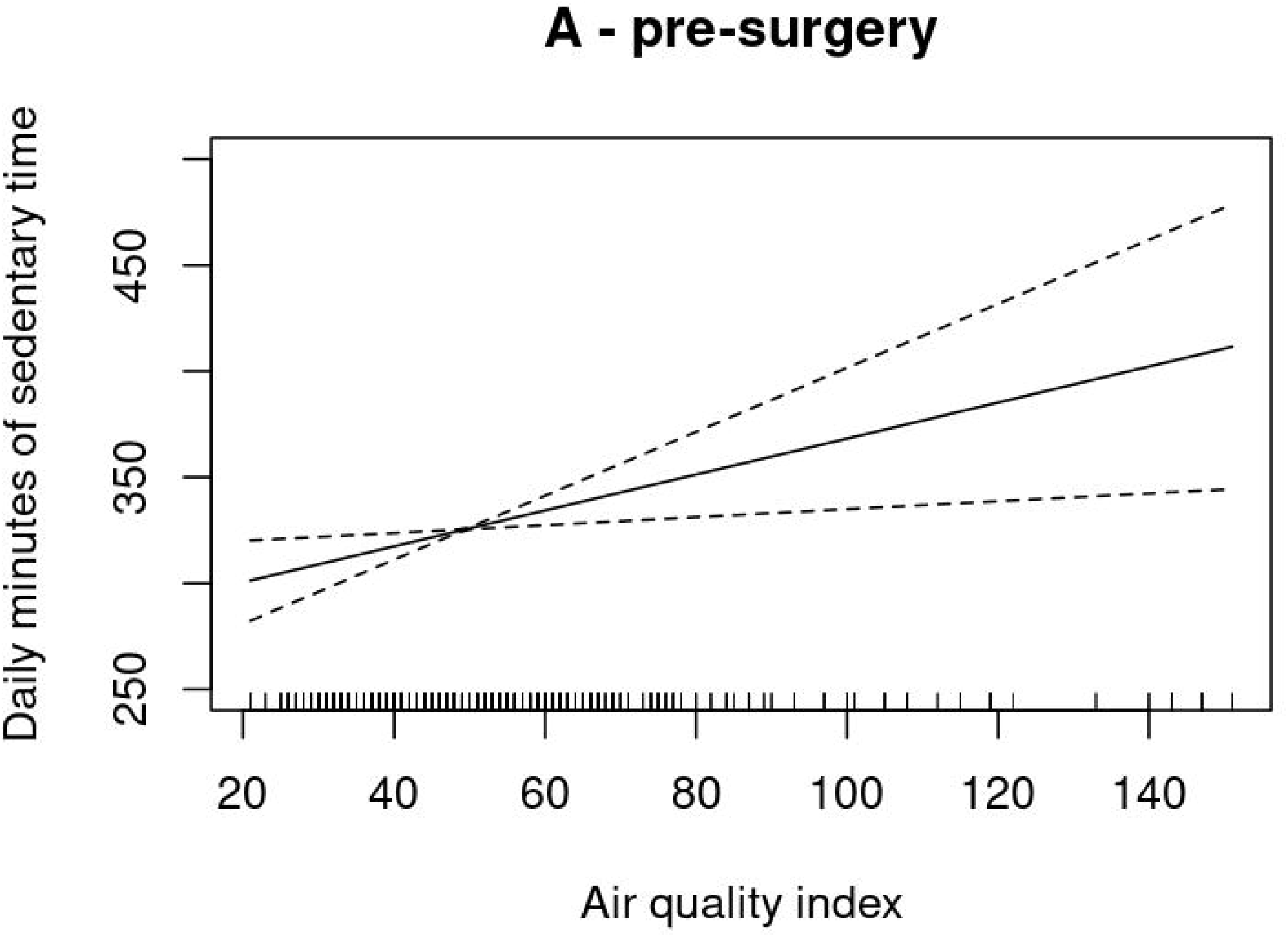
Linear association of air quality index with sedentary time before MBS.

## Discussion

MBS patients are encouraged to engage in regular physical activity to optimize and sustain postoperative weight loss and health improvements. However, many patients are insufficiently active before MBS and struggle to achieve and/or maintain sufficient physical activity levels after MBS. This warrants greater attention to identifying and understanding barriers to physical activity in this patient population. To date, efforts in this regard have mostly focused on individual-level barriers with minimal consideration of how patients’ external environment might inhibit physical activity. Although some data suggest that weather could be a barrier for some MBS patients, no study has systematically evaluated associations of specific weather and air pollution indices with physical activity and sedentary behaviors in the context of MBS. This may be particularly important given ongoing and future climate changes (e.g., higher temperatures) and obesity-related vulnerabilities (e.g., diminished heat tolerance during physical activity) to these changes. Therefore, this study aimed to improve understanding of how variability in objective temperature and air pollution indices relate to objectively-measured physical activity and sedentary behaviors on a daily level among adults with obesity before and during the initial year after MBS.

Our results showed that MVPA related in a nonlinear fashion to temperature indices. Regarding maximal outdoor temperatures, a large decrease in daily MVPA was observed for maximal temperatures exceeding ∼20°C (∼68°F). The only study found in adults with obesity also showed an inverse U-shape with the probability of monthly leisure-time physical activity participation markedly decreasing after 36 °C^19^.

Similarly, for WBGT, which considers humidity, wind speed, sun angle, and cloud cover along with temperature, the highest MVPA levels were observed between ∼ - 7°C (∼20°F) and ∼14°C (∼57°F) WBGT, with a drastic reduction of MVPA below and above these values. This WBGT level falls well below the WBGT level of > 25.7°C (78.1°F) that is associated with high risk to perform MVPA for unfit or non-acclimatized adults. Thus, our findings align with previous research suggesting that higher temperatures have a negative effect on physical activity of higher intensities among adults and children with obesity^35,36^.

Regarding MVPA sensitivity analyses, although overall associations of MVPA and temperature indices were similar between pre- and post-MBS, visual inspection of pre- and post-MBS plots displayed a less marked decrease of daily MVPA minutes after MBS for average temperature of ∼20°C (∼68°F) and WBGT of ∼8°C (∼47°F) compared to before MBS. Although additional research is needed to substantiate this observation, it is possible that postoperative reduction in subcutaneous fat could contribute to improved capacity to acclimate to warmer temperatures when performing MVPA. Indeed, higher levels of subcutaneous fat impede heat loss contributing to lower capacity to acclimate while performing MVPA in warmer temperatures increasing risk of heat stress^37^.

In accordance with studies using self-reported MVPA and other physical activity indicators (total volume, daily step counts, leisure)^13,14,38^, our results also showed that daily MVPA is negatively associated with the level of air pollution. Specifically, MVPA levels decreased after ∼70 AQI score. Although this value is considered acceptable for the general population, it is considered a risk for some people who are unusually sensitive to air pollution^28^. People with obesity may be more sensitive to air pollutants during MVPA than individuals without obesity given the inflammatory and mechanical (i.e., excess fat deposited on the chest wall and abdominal cavity) effects of obesity decreasing respiratory function and exercise tolerance^39,40^. We also found that time spent in sedentary behaviors before surgery increased with higher levels of air pollution. Taken together, these findings suggest that higher levels of air pollution may contribute to low MVPA among MBS patients, and increased sedentary time especially before MBS.

The above findings may have important clinical implications for prescribing and promoting physical activity among MBS patients. For example, during seasons with warmer temperatures, patients might be encouraged to perform MVPA during the morning hours to limit exposure to hotter temperatures. When exercising outside, patients should be encouraged to hydrate before, during and after their session, wear light-weight clothing, and use wearable cooling wraps (e.g., around neck) to enhance thermoregulatory capacity. Patients might also benefit from exercise outside and before 9 AM given that air pollution tends to be higher in homes and workplaces than outside and peaks at 9 AM on average^41,42^. More broadly, it suggests that coordinated effort to develop cooling strategies at urban, building and individual scales will be particularly beneficial for MBS patients^43^.

As more and more evidence showed that LPA is related to health benefits in adults^44^, this study also explored weather indices, and air pollution associations with LPA. Our results showed no statistically significant association of LPA with weather indices, and air pollution, in contrast with some studies reporting association between LPA and outdoor temperature among older population^7,45^. Nevertheless, evidence on the association between LPA and outdoor temperature is inconsistent, and sparse for air quality^7-9^. Additional studies are therefore required, and could also consider the different forms of LPA (leisure, transport-related or occupational activity), that might impact the relationship between outdoor environmental factors and LPA.

As regard to daily time spent in sedentary behaviors, no statistically significant association was also found with temperature and WBGT in the current study. Our results on outdoor temperature are consistent with some studies, but not with others^7-9^. In addition, our results showed positive associations between ST and AQI before MBS, but not after MBS. These discrepancies between all the studies may be explained by several factors, such as different in populations, regions, climatic zones studied, statistical model used or covariates considered. Additional studies are required to disentangle discordances, and explore confounding potential factors. For example, the facts that sedentary behaviors are usually indoor activities, the impact of outdoor environmental factors on them could be minimal, especially in MBS adults spending on average more than 80% of their time in sedentary behaviors^3^.

## Strengths and limits

To our knowledge, this study is the first to prospectively explore the associations of objective weather and air quality indices with objectively measured physical activity and sedentary time on a daily level before and after MBS. By using a multilevel non-linear statistical modelling, an approach too rarely used in physical activity domain^7,31^, our study allows to model nonlinear relationships. Nevertheless, several limitations of this study must also be acknowledged. First, the generalization of our results is limited to temperate climatic regions, and cities with rare episode of unhealthy air quality. Second, the accelerometer did not include GPS data, and participant’s address zip codes were not collected, making impossible to know the weather station closest to the daily location of participants. Then, other potential environmental factors not assessed could have influenced the associations (e.g. cooling strategies availability or walkability neighborhoods). Finally, accelerometers do not provide information on the context (e.g. indoor or outdoor), and the form (leisure, transport-related or occupational activity) of physical activity and sedentary behavior. Thus, it is possible that stronger effects might be observed if all PA was performed outdoors.

## Conclusion

Rising average global temperatures and related widespread worsening of changes in weather patterns and air pollution pose significant threats to engagement in important health behaviors such as physical activity. Physical activity levels of individuals with obesity may be especially affected by these environmental impacts given that obesity is associated with factors such as vulnerability to heat stress and impaired respiratory strength that make performing commonly prescribed aerobic activities such as walking more difficult. The current study is the first to show that higher levels of temperature and air pollution indices contribute to lower MVPA and higher ST in the context of MBS. Less marked decreases in MVPA at similar higher temperatures after MBS compared to before MBS suggests patients may have increased heat tolerance during MVPA although confirmation of this finding and mechanisms is needed. Weather and environmental factors should be considered in prescriptions and strategies to promote more active lifestyles among MBS patients.

## Data Availability

Data described in the manuscript, and code book will be made available on request pending application. The R syntax for main analyses are provided on the Open Sciences Framework (https://osf.io/34usq/).

## Acknowledgments

We would like to thank Jennifer Webster for her assistance in executing the study procedures, and the study participants for their commitment to this study. Data described in the manuscript, and code book will be made available on request pending application. The R syntax for main analyses are provided on the Open Sciences Framework (https://osf.io/34usq/).

## Author contributions

Dale S. Bond, J. Graham Thomas, Aurelie Baillot and Jmii, Nejm Eddine acquired the data. Aurelie Baillot, Paquito Bernard, and Dale S. Bond conceived the study and design. Paquito Bernard performed statistical analysis. Aurelie Baillot, Paquito Bernard and Dale S. Bond drafted the manuscript. All authors revised the manuscript for intellectual content and approved the final version of the completed manuscript.

CLINICAL TRIAL REGISTRATION: NCT02777177

FUNDING: R01 DK108579 (MPI: Bond and Thomas)

## References

1. Bellicha A, van Baak MA, Battista F, Beaulieu K, Blundell JE, Busetto L, et al. Effect of exercise training before and after bariatric surgery: A systematic review and meta-analysis. Obes Rev. 2021;22 Suppl 4:e13296.doi: 10.1111/obr.13296.

2. Nielsen MS, Alsaoodi H, Hjorth MF, Sjodin A. Physical Activity, Sedentary Behavior, and Sleep Before and After Bariatric Surgery and Associations with Weight Loss Outcome. Obes Surg. 2021;31(1):250–9.doi: 10.1007/s11695-020-04908-3.

3. Bond DS, Unick JL, Jakicic JM, Vithiananthan S, Pohl D, Roye GD, et al. Objective assessment of time spent being sedentary in bariatric surgery candidates. Obes Surg. 2011;21(6):811–4.doi: 10.1016/j.soard.2008.08.003.

4. King WC, Bond DS. The importance of pre and postoperative physical activity counseling in bariatric surgery. Exerc Sport Sci Rev. 2013;41(1):26–35.doi: 10.1097/JES.0b013e31826444e0.

5. Sallis JF, Cervero RB, Ascher W, Henderson KA, Kraft MK, Kerr J. An ecological approach to creating active living communities. Annu Rev Public Health. 2006;27:297–322.doi: 10.1146/annurev.publhealth.27.021405.102100.

6. Bauman AE, Reis RS, Sallis JF, Wells JC, Loos RJ, Martin BW. Correlates of physical activity: why are some people physically active and others not? Lancet. 2012;380(9838):258–71.doi: 10.1016/S0140-6736(12)60735-1.

7. Turrisi TB, Bittel KM, West AB, Hojjatinia S, Hojjatinia S, Mama SK, et al. Seasons, weather, and device-measured movement behaviors: a scoping review from 2006 to 2020. Int J Behav Nut Phys Act. 2021;18(1):24.doi: 10.1186/s12966-021-01091-1.

8. Zisis E, Hakimi S, Lee EY. Climate change, 24-hour movement behaviors, and health: a mini umbrella review. Glob Health Res Policy. 2021;6(1):15.doi: 10.1186/s41256-021-00198-z.

9. Bernard P, Chevance G, Kingsbury C, Baillot A, Romain AJ, Molinier V, et al. Climate Change, Physical Activity and Sport: A Systematic Review. Sports Med. 2021;51(5):1041–59.doi: 10.1007/s40279-021-01439-4.

10. Al-Mohannadi AS, Farooq A, Burnett A, Van Der Walt M, Al-Kuwari MG. Impact of Climatic Conditions on Physical Activity: A 2-Year Cohort Study in the Arabian Gulf Region. J Phys Act Health. 2016;13(9):929–37.doi: 10.1123/jpah.2015-0593.

11. Ahn Y, Okamoto D, Uejio C. Investigating city bike rental usage and wet-bulb globe temperature. Int J Biometeorol. 2022;66(4):679–90.doi: 10.1007/s00484-021-02227-5.

12. Mix JM, Elon L, Thein Mac VV, Flocks J, Economos J, Tovar-Aguilar AJ, et al. Physical activity and work activities in Florida agricultural workers. Am J Ind Med. 2019;62(12):1058–67.doi: 10.1002/ajim.23035.

13. An R, Zhang S, Ji M, Guan C. Impact of ambient air pollution on physical activity among adults: a systematic review and meta-analysis. Perspect Public Health. 2018;138(2):111–21.doi: 10.1177/1757913917726567.

14. Roberts JD, Voss JD, Knight B. The association of ambient air pollution and physical inactivity in the United States. PLoS One. 2014;9(3):e90143.doi: 10.1371/journal.pone.0090143.

15. Yu H, Cheng J, Gordon SP, An R, Yu M, Chen X, et al. Impact of Air Pollution on Sedentary Behavior: A Cohort Study of Freshmen at a University in Beijing, China. Int J Environ Res Public Health. 2018;15(12).doi: 10.3390/ijerph15122811.

16. Baillot A, Chenail S, Barros Polita N, Simoneau M, Libourel M, Nazon E, et al. Physical activity motives, barriers, and preferences in people with obesity: A systematic review. PLoS One. 2021;16(6):e0253114.doi: 10.1371/journal.pone.0253114.

17. Bond DS, Thomas JG, Ryder BA, Vithiananthan S, Pohl D, Wing RR. Ecological momentary assessment of the relationship between intention and physical activity behavior in bariatric surgery patients. Int J Behav Med. 2013;20(1):82–7.doi: 10.1007/s12529-011-9214-1.

18. Chan CB, Ryan DA, Tudor-Locke C. Relationship between objective measures of physical activity and weather: a longitudinal study. Int J Behav Nut Phys Act. 2006;3:21.doi: 10.1186/1479-5868-3-21.

19. Obradovich N, Fowler J H. Climate Change May Alter Human Physical Activity Patterns. Nat Hum Behav. 2017;1(5):0097.

20. United States Environmental protection Agency. Technical Documentation: Heat Waves 2021 [2022-11-10]. Available from: https://www.epa.gov/sites/default/files/2021-04/documents/heat-waves_td.pdf.

21. IPCC. Climate Change 2021: The Physical Science Basis. Contribution of Working Group I to the Sixth Assessment Report of the Intergovernmental Panel on Climate Change. Cambridge, United Kingdom and New York, NY, USA: 2021.

22. Chevance G, Fresan U, Hekler E, Edmondson D, Lloyd SJ, Ballester J, et al. Thinking Health-related Behaviors in a Climate Change Context: A Narrative Review. Ann Behav Med. 2022.doi: 10.1093/abm/kaac039.

23. Goldstein SP, Thomas JG, Vithiananthan S, Blackburn GA, Jones DB, Webster J, et al. Multi-sensor ecological momentary assessment of behavioral and psychosocial predictors of weight loss following bariatric surgery: study protocol for a multicenter prospective longitudinal evaluation. BMC Obes. 2018;5:27.doi: 10.1186/s40608-018-0204-6.

24. Cole RJ, Kripke DF, Gruen W, Mullaney DJ, Gillin JC. Automatic sleep/wake identification from wrist activity. Sleep. 1992;15(5):461–9.doi: 10.1093/sleep/15.5.461.

25. Tudor-Locke C, Barreira TV, Schuna JM, Jr., Mire EF, Katzmarzyk PT. Fully automated waist-worn accelerometer algorithm for detecting children’s sleep-period time separate from 24-h physical activity or sedentary behaviors. Appl Physiol Nutr Metab. 2014;39(1):53–7.doi: 10.1139/apnm-2013-0173.

26. Kamada M, Shiroma EJ, Harris TB, Lee IM. Comparison of physical activity assessed using hip- and wrist-worn accelerometers. Gait Posture. 2016;44:23–8.doi: 10.1016/j.gaitpost.2015.11.005.

27. National Weather Service. Air Quality Index [cited 2023 2023-02-17]. Available from: https://www.weather.gov/safety/airquality-aqindex#:~:text=The%20Air%20Quality%20Index%20(AQI,days%20after%20breathing%20polluted%20air.

28. Cromar KR, Ghazipura M, Gladson LA, Perlmutt L. Evaluating the U.S. Air Quality Index as a risk communication tool: Comparing associations of index values with respiratory morbidity among adults in California. PLoS One. 2020;15(11):e0242031.doi: 10.1371/journal.pone.0242031.

29. American College of Sports M, Armstrong LE, Casa DJ, Millard-Stafford M, Moran DS, Pyne SW, et al. American College of Sports Medicine position stand. Exertional heat illness during training and competition. Med Sci Sports Exerc. 2007;39(3):556–72.doi: 10.1249/MSS.0b013e31802fa199.

30. Ravindra K, Rattan P, Mor S, Aggarwal AN. Generalized additive models: Building evidence of air pollution, climate change and human health. Environ Int. 2019;132:104987.doi: 10.1016/j.envint.2019.104987.

31. Bernard P, Dore I, Romain AJ, Hains-Monfette G, Kingsbury C, Sabiston C. Dose response association of objective physical activity with mental health in a representative national sample of adults: A cross-sectional study. PLoS One. 2018;13(10):e0204682.doi: 10.1371/journal.pone.0204682.

32. Wood S. Generalized additive models: an introduction with R. Raton B, editor: FL: Chapman & Hall/CRC; 2006.

33. Vrieze SI. Model selection and psychological theory: a discussion of the differences between the Akaike information criterion (AIC) and the Bayesian information criterion (BIC). Psychol Methods. 2012;17(2):228–43.doi: 10.1037/a0027127.

34. Giorgini P, Rubenfire M, Das R, Gracik T, Wang L, Morishita M, et al. Higher fine particulate matter and temperature levels impair exercise capacity in cardiac patients. Heart. 2015;101(16):1293–301.doi: 10.1136/heartjnl-2014-306993.

35. Foster J, Hodder SG, Lloyd AB, Havenith G. Individual Responses to Heat Stress: Implications for Hyperthermia and Physical Work Capacity. Front Physiol. 2020;11:541483.doi: 10.3389/fphys.2020.541483.

36. Dougherty KA, Chow M, Kenney WL. Responses of lean and obese boys to repeated summer exercise in the heat bouts. Med Sci Sports Exerc. 2009;41(2):279–89.doi: 10.1249/MSS.0b013e318185d341.

37. Speakman JR. Obesity and thermoregulation. Handb Clin Neurol. 2018;156:431–43.doi: 10.1016/B978-0-444-63912-7.00026-6.

38. Yu H, Yu M, Gordon SP, Zhang R. The association between ambient fine particulate air pollution and physical activity: a cohort study of university students living in Beijing. Int J Behav Nutr Phys Act. 2017;14(1):136.doi: 10.1186/s12966-017-0592-x.

39. Dixon AE, Peters U. The effect of obesity on lung function. Expert Rev Respir Med. 2018;12(9):755–67.doi: 10.1080/17476348.2018.1506331.

40. McNeill JN, Lau ES, Zern EK, Nayor M, Malhotra R, Liu EE, et al. Association of obesity-related inflammatory pathways with lung function and exercise capacity. Respir Med. 2021;183:106434.doi: 10.1016/j.rmed.2021.106434.

41. Cichowicz R, Stelegowski A. Average Hourly Concentrations of Air Contaminants in Selected Urban, Town, and Rural Sites. Arch Environ Contam Toxicol. 2019;77(2):197–213.doi: 10.1007/s00244-019-00627-8.

42. Roh T, Moreno-Rangel A, Baek J, Obeng A, Hasan NT, Carrillo G. Indoor Air Quality and Health Outcomes in Employees Working from Home during the COVID-19 Pandemic: A Pilot Study. Atmosphere. 2021;12:1665.

43. Jay O, Capon A, Berry P, Broderick C, de Dear R, Havenith G, et al. Reducing the health effects of hot weather and heat extremes: from personal cooling strategies to green cities. Lancet. 2021;398(10301):709–24.doi: 10.1016/S0140-6736(21)01209-5.

44. Amagasa S, Machida M, Fukushima N, Kikuchi H, Takamiya T, Odagiri Y, et al. Is objectively measured light-intensity physical activity associated with health outcomes after adjustment for moderate-to-vigorous physical activity in adults? A systematic review. Int J Behav Nutr Phys Act. 2018;15(1):65.doi: 10.1186/s12966-018-0695-z.

45. Davis MG, Fox KR, Hillsdon M, Sharp DJ, Coulson JC, Thompson JL. Objectively measured physical activity in a diverse sample of older urban UK adults. Med Sci Sports Exerc. 2011;43(4):647–54.doi: 10.1249/MSS.0b013e3181f36196.

